# A low-cost culture- and DNA extraction-free method for the molecular detection of pneumococcal carriage in saliva

**DOI:** 10.1101/2023.12.07.23299679

**Authors:** Chikondi Peno, Tzu-Yi Lin, Maikel S. Hislop, Devyn Yolda-Carr, Katherine Farjado, Anna York, Virginia E. Pitzer, Daniel M. Weinberger, Amy K. Bei, Orchid M. Allicock, Anne L. Wyllie

**Author notes:** Correspondence: Anne Wyllie, 60 College St, New Haven, CT 06510, Chikondi Peno, 60 College St, New Haven, CT 06510. These authors contributed equally (Chikondi Peno, Tzu-Yi Lin).

## Abstract

**Background:** Molecular methods have improved the sensitivity of detection of pneumococcal carriage in saliva. However, they typically require sample culture-enrichment and nucleic acid extraction, prior to performing the detection assay. These factors may limit scalability for extensive surveillance of pneumococcus, particularly in low-resource settings. In this study, we evaluated the performance of a DNA-extraction-free method for the detection of pneumococcus in saliva.

**Methods:** We developed a streamlined qPCR-based protocol for the detection of pneumococcus, omitting culture-enrichment and DNA extraction. Using saliva samples collected from children attending childcare centers (New Haven, CT, USA), we evaluated detection of pneumococcus using saliva lysates as compared to purified DNA extracted from culture-enriched aliquots of the paired samples using qPCR targeting the pneumococcal *piaB* gene.

**Results:** Of 759 saliva samples tested from 92 children (median age 3.65 years; IQR (2.46-4.78), pneumococcus was detected in 358 (47.2%) saliva lysates prepared using the extraction-free protocol and in 369 (48.6%) DNA extracted from the culture-enriched samples. We observed a near-perfect agreement between the two protocols (Cohen’s kappa: 0.92; 95%CI: 0.90-0.95). While we also observed a high correlation between the qPCR C_T_ values generated by the two methods (r=0.93, *p*<0.0001), the C_*T*_ values generated from the extraction-free, saliva lysates were higher (lower concentration) than those obtained from DNA extracted from culture-enriched samples (ΔC_*T*_ = 6.68, *p*<0.00001).

**Conclusions:** For pneumococcal carriage surveillance in children, our findings suggest that a DNA extraction-free approach may offer a cost-effective alternative to the resource-intensive culture-enrichment method. While, as expected, we observed higher qPCR C_*T*_ values (lower bacterial load) in the absence of culture-enrichment, the overall rate of detection remained unaffected.

## BACKGROUND

*Streptococcus pneumoniae* (pneumococcus) is a leading cause of bacterial infections including pneumonia, meningitis, bacteremia and sepsis. Globally, more than 300,000 deaths in children under the age of five are caused by pneumococcal diseases, with the highest burden of disease experienced in low-to-middle income countries.^1^ Carriage of pneumococcus in the upper respiratory tract (URT) is common and usually asymptomatic, but also a prerequisite for invasive pneumococcal disease and a source of pneumococcal transmission in the community.^2,3^ Following the introduction of pneumococcal vaccines, carriage surveillance has been pivotal for evaluating vaccine performance and informing new vaccination strategies.^4,5^ Because pneumococcal vaccines only target a subset of >100 known pneumococcal serotypes,^6^ carriage surveillance is also crucial for monitoring persisting vaccine serotypes and detecting emergence and expansion of non-vaccine serotypes that can be targeted in new vaccine formulations.^7^

The surveillance of pneumococcal carriage dynamics relies on establishing methods of detection that are sufficiently sensitive across all ages and scalable in different settings, particularly in settings with a high disease burden.^8^ While the current recommended gold standard method for detecting pneumococcal carriage in the URT is conventional culture of a nasopharyngeal swab,^9^ molecular detection methods improve the sensitivity of pneumococcal carriage detection when applied to nasopharyngeal swabs^10,11^ and to other URT sample types including oropharyngeal swabs^12^ and saliva.^3,11,13^ Despite being more sensitive for pneumococcal carriage detection, molecular methods are resource-intensive, requiring extraction of DNA from the sample prior to testing. Moreover, for oral sample types (oropharyngeal swabs and saliva), a culture step using selective medium is recommended prior to nucleic acid extraction to enrich the sample for pneumococcus to further improve the sensitivity of detection in these polymicrobial samples.^14^ Together, these requirements may limit scalability for extensive surveillance efforts, particularly in resource-limited settings. Saliva, however, is an ideal sample for pneumococcal community surveillance studies; it is a non-invasive sample type which can be reliably self-collected,^15^ negating the need for trained healthcare professionals, typically required for the collection of swabs. Overall, this leads to lower sample collection burden to study participants (minimizing potential testing aversion), and/or clinical personnel, which is critical particularly in longitudinal studies. Additionally, we have previously demonstrated that encapsulated pneumococci remain viable in raw, unsupplemented saliva with a stable bacterial load seen for 24 hours in the absence of cold-chain transport (i.e., ∼19-30°C),^16^ further alleviating collection and transport burden for studies conducted in remote or resource-limited settings.

During the COVID-19 pandemic, Vogels *et al*. developed a saliva-based, nucleic acid extraction-free qPCR test for the detection of SARS-CoV-2 with the aim of reducing the burden and cost of molecular detection.^17^ Replacing nucleic acid extraction with a simple enzymatic lysis or heat treatment step was possible without compromising the accuracy and efficiency of testing for SARS-CoV-2.^17^ Here, we evaluated the application of this extraction-free qPCR approach to detect pneumococcus in saliva samples, to explore the feasibility of a low-cost, molecular screening method screening tool for pneumococcal carriage.

## METHODS

### Ethics

The collection of saliva samples from children in childcare centers (New Haven, CT, USA) was approved by the Institutional Review Board of the Yale Human Research Protection Program (Protocol number: 200002839). Written informed consent was obtained from parent or guardian of every participating child. The collection of de-identified saliva samples from healthy volunteers for assay validation was approved by the Institutional Review Board of the Yale Human Research Protection Program (Protocol number: 2000029374).

### Study design

We compared methods for the detection of carriage of the pneumococcus in saliva samples collected weekly from children to screen for the prevalence of SARS-CoV-2 in New Haven childcare centers over the 2020/2021 and 2021/2022 school years. Methods used for enrolment of study participants and at-home saliva collection have been previously described.^18^ After testing for SARS-CoV-2, remaining sample volumes were stored at -80°C until further analysis. Due to sample handling requirements during the COVID-19 pandemic, for samples collected during the 2020/2021 school year, unsupplemented raw saliva was stored and used for the detection of pneumococcus. For samples collected during 2021/2022 school year, saliva aliquots for the detection of pneumococcus using the culture enrichment method were supplemented with Brain Heart Infusion (BHI; Oxoid) supplemented with 10% glycerol, while the remaining aliquot tested using the extraction-free method was stored as unsupplemented raw saliva.

### Standard culture-enrichment and DNA extraction protocol for pneumococcal carriage detection

Samples were thawed on ice, and 100 μL of raw saliva was inoculated on trypticase soy agar (TSA II) supplemented with 5% defibrinated sheep blood and 10 mg/L gentamicin (Gent plates, ThermoFisher Scientific) and incubated overnight at 37°C with 5% CO_2._^19^ Bacterial growth was harvested into 2,100 μL of BHI supplemented with 10% glycerol and stored at -80°C until further processing. These samples were considered as culture-enriched for pneumococcus. DNA was extracted from 200 μL of culture-enriched samples using the MagMAX Viral/Pathogen Nucleic Acid Isolation kit (ThermoFisher Scientific), following a modified protocol on the KingFisher Apex (ThermoFisher Scientific) as previously described.^20^

### Extraction-free protocol

Saliva samples were thawed on ice, and lysates were prepared following the “SalivaDirect” extraction-free protocol as previously described (Figure 1).^17^ Briefly, a total of 6.5 µL (20 mg/mL) of proteinase K was added to 50 µL of each saliva sample, aliquoted into strip tubes. The tubes were firmly sealed, placed in a rack, and vortexed for 1 minute at 3200 RPM. Samples were then heated on a thermocycler at 95°C for 5 minutes.

**Figure 1.**
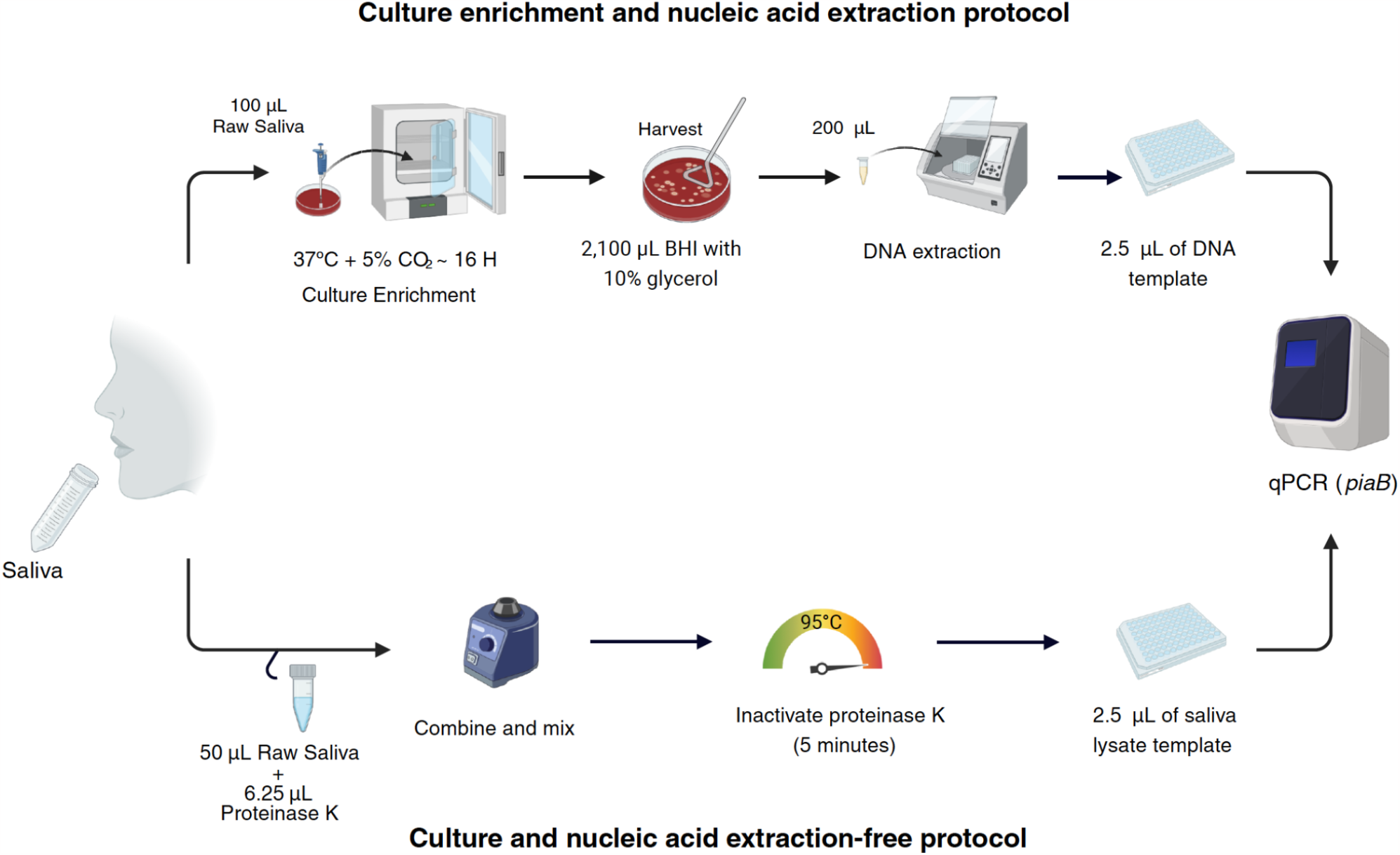
Schematic overview of the sample preparation methods compared for the detection of pneumococcal carriage in saliva samples. Raw, unsupplemented saliva was tested using qPCR targeting the pneumococcus-specific gene, piaB, following A) culture-enrichment on trypticase soy agar supplemented with 5% defibrinated sheep blood and 10 mg/L gentamicin (Gent plates, ThermoFisher Scientific) prior to DNA extraction, and B) a simplified culture- and extraction-free approach in which saliva was simply mixed with proteinase K and heated at 95°C for 5 minutes. Figure created with Biorender.com.

### Molecular detection of pneumococcal carriage

A real-time qPCR assay targeting the pneumococcal iron uptake ABC transporter lipoprotein *piaB* gene was used to detect the presence of pneumococcal DNA in both the DNA extracted from culture-enriched samples and saliva lysates, as previously described.^12,19,20^ Both DNA extracts and saliva lysates (2.5 µL) were tested in 20 µL reaction volumes using Advanced Universal Probe Supermix (BioRad, USA) and a primer/probe mix at concentrations of 250 nM (1 μL per reaction) (Iowa Black quenchers, USA). A standard curve prepared from five 1:10 serial dilutions of pneumococcal DNA, ranging from 1 ng/μL to 0·00001 ng/μL was included as a positive control. Water was added as a non-template/negative qPCR control in each run. In qPCR runs where extracted DNA was used as a template, a negative extraction control (BHI+10% Glycerol) was included. Samples were classified as positive for pneumococcus when a *piaB* cycle threshold (C_*T*_) value was <40.^19,20^

### Limit of detection of the extraction-free approach

Saliva samples were collected from healthy volunteers and screened for the absence of pneumococcus using qPCR targeting *piaB*, as previously described.^16^ Samples negative for both targets were considered negative for pneumococcus and were pooled together. A serotype 19A pneumococcus strain was serially diluted into the pooled saliva, from 5X10^7^ CFU/ml to 5X10^1^ CFU/mL, in duplicate (Supplementary Figure 1).^21^ Each dilution was tested in triplicate, using both the extraction-free method and culture-enrichment methods as described above. The limit of detection was determined as the bacterial concentration in which all the triplicates in both replicates were positive for pneumococcus (C_*T*_<40).

### Statistical analysis

Cohen’s kappa was used to assess the chance-corrected agreement of identifying pneumococcal carriage based on the two methods.^22^ Cohen’s kappa ratios of 0, 0.01–0.20, 0.21–0.40, 0.41–0.60, 0.61–0.80, and >0.81 were interpreted as exhibiting poor, slight, fair, moderate, substantial, and near-perfect agreement, respectively.^22,23^ In addition, McNemar’s test was used to assess the discordance between the two methods, to complement Cohen’s Kappa. Screening test parameters (sensitivity, specificity, positive and negative predictive values) were calculated using the “caret” R package by using the culture-enrichment protocol or composite reference as the “gold standard”, where pneumococcal positivity was defined as being positive by either culture-enrichment method or the extraction-free method.^24^ We used linear regression to evaluate the differences in C_*T*_ values obtained using the two-methods. Because samples collected between the two sampling periods were stored differently (i.e., without supplementation in 2020/2021 school year and supplemented with BHI+10% Glycerol in 2021/2022 school year) before culture-enrichment was performed, we fitted a linear regression model adjusting for the sampling period to take into account differences in storage conditions between the two sampling periods. An interaction term between method and sampling period was also fitted to evaluate whether the effect of the method varied by sampling period. p-values of less than 0.05 were considered significant. All analyses were performed using R (version 4.3.0).

## RESULTS

### Performance of the extraction-free method in the detection of pneumococcal carriage

A total of 759 saliva samples collected from 92 children attending child care centers (median age: 3.23 years, IQR: 1.8-4.5 years) were used to compare the performance of two pneumococcal detection protocols: a well-established method in which DNA is extracted following sample culture-enrichment^3,13,25,26^ and an extraction-free method, originally developed and extensively used for the detection of SARS-CoV-2 (Figure 1).^17^ Of the 759 samples, pneumococcus was detected in 378 (49.8%) samples using at least one of the methods, with a near-perfect agreement between the two protocols (Cohen’s kappa=0.92, 95%CI: 0.90-0.95; Table 1) the culture-enrichment protocol detected slightly more positive samples (369/759, 48.6%) compared to the extraction-free method (358/759, 47.1%). We observed discordant results in 29/378 (7.6%) samples, of which 20/29 (68.9%) tested positive using the culture-enrichment method but negative by the extraction-free protocol, while 9/29 (31.0%) tested positive using the extraction-free method but negative following culture-enrichment (Figure 2A). There was no significant difference in the detection rate of the two methods according to McNemar’s test (*p*=0.06). Almost all samples that yielded discordant results (28/29, 96.5%) had higher qPCR C_*T*_ values (C_*T*_ >30), with the majority having C_*T*_ values >35 (23/29; 79.3%) (Supplementary Table 1).

**Table 1.**
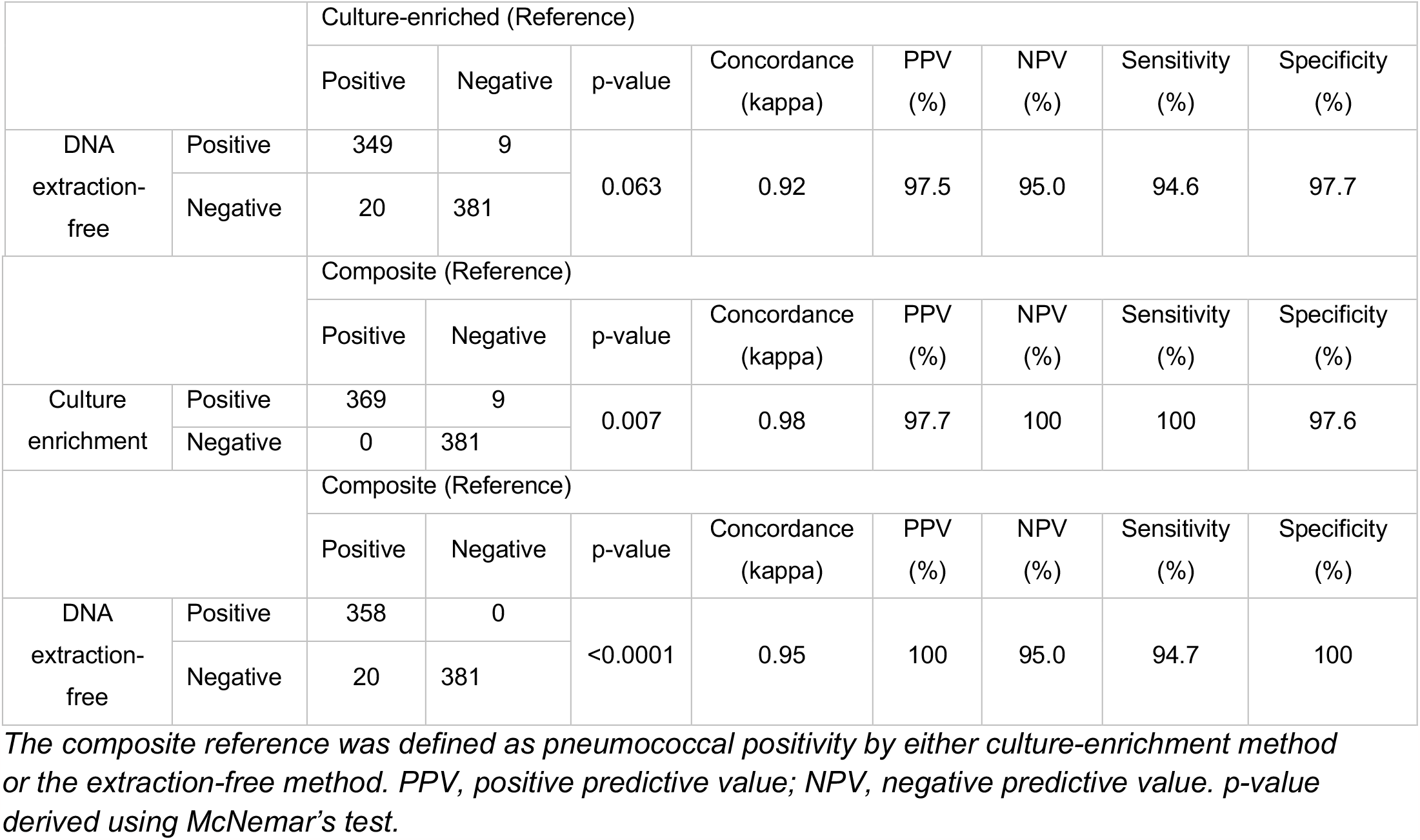
Sensitivity and specificity of pneumococcal carriage detection in saliva samples when tested using qPCR following culture-enrichment or extraction-free sample processing methods.

**Figure 2.**
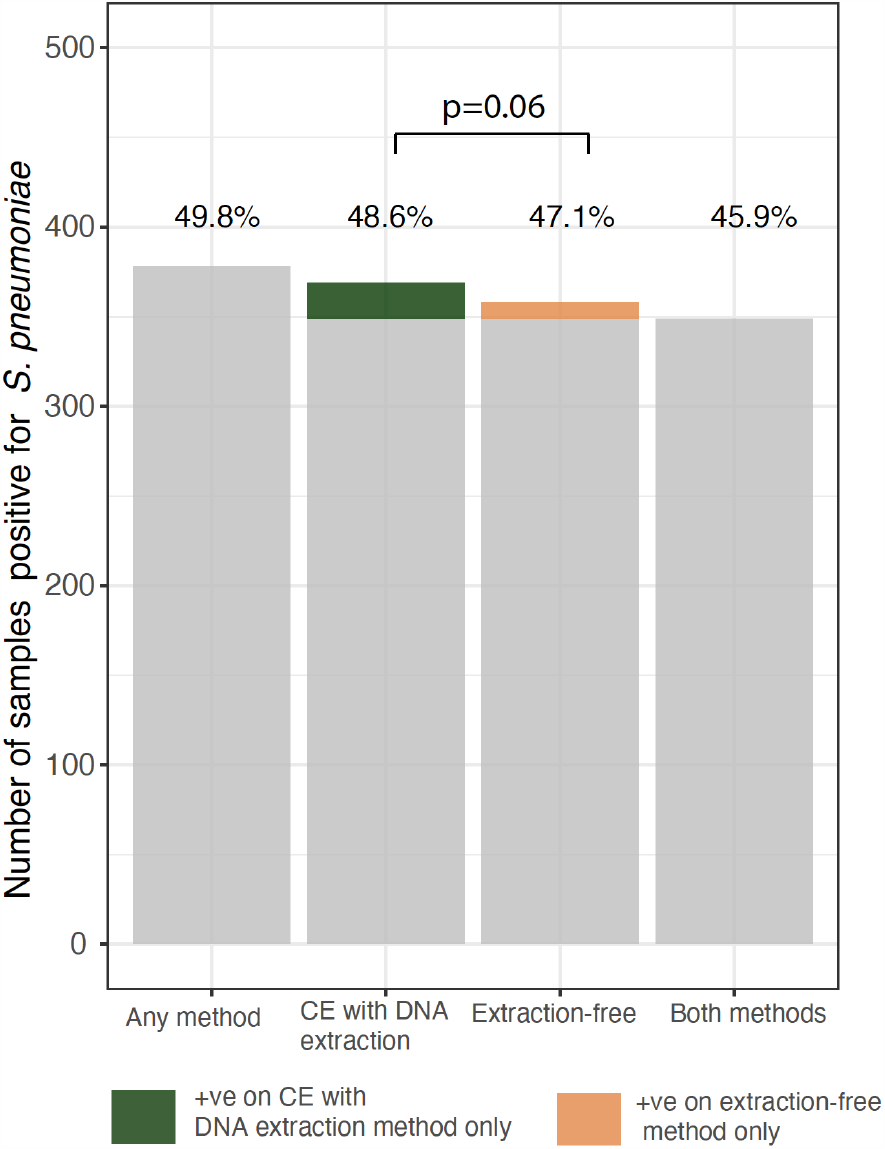
Detection of pneumococcus in raw, unsupplemented saliva using methods with and without sample culture-enrichment and DNA extraction. Overall, 378/759 (49.8%) saliva samples tested positive for pneumococcus (piaB qPCR C_T_ value <40), when tested by either culture-enrichment followed by DNA extraction (“CE with DNA extraction”) and a culture- and extraction-free method (“Extraction-free”). Of these, 369/759 (48.6%) tested positive using the CE with DNA extraction method compared to 358/759 (47.1%) when using the extraction-free method; 348/759 (45.9%) tested positive in both methods. The proportion of samples found to be positive using the CE with DNA extraction method highlighted in green denote those that tested positive using this method only (n=20). The proportion of samples found to be positive using the extraction-free method highlighted in orange denote samples that tested positive when using this method only (n=9).

The accuracy of detection when testing samples using the extraction-free method was not inferior to that of the culture-enrichment method. When using the culture-enrichment method as the “gold standard,” all measures of accuracy including sensitivity (94.7%), specificity (97.7%), positive predictive value (97.5%) and negative predictive value (95%) were greater than 90% (Table 1). Since the culture-enrichment method is not an established gold standard, we further compared the accuracy of the detection methods using a composite reference, where pneumococcal carriage positivity was defined as any sample yielding a C_*T*_ value <40 from either the culture-enrichment or the extraction-free method. For both methods there was a near-perfect agreement with the composite reference (culture-enrichment, Cohen’s kappa=0.98 vs. extraction-free, Cohen’s kappa=0.95; Table 1). However, there was significant disagreement for both methods when compared to the composite reference (culture-enrichment, McNemar’s test *p*=0.007 vs. extraction-free, Mcnemar’s test *p*<0.0001; Table 1). Nonetheless, all measures of detection accuracy remained above 90% (Table 1).

The limit of detection of pneumococcus when testing saliva lysates processed by the extraction-free method was determined to be 5X10^3^ CFU/mL. When testing DNA extracted from culture-enriched samples, we were able to detect pneumococcus up to 5X10^1^ CFU/mL (Figure S3), demonstrating the lower quantification capacity of the extraction-free method when compared to the culture-enrichment method.

We observed a high correlation between the C_*T*_ values obtained using the extraction-free method and the culture-enrichment method (r=0.92, *p*<0.00001; Figure 3A). Among the samples yielding positive results on both methods, lower bacterial quantities (as inferred by higher qPCR C_*T*_ values) were observed when using the extraction-free protocol as compared to the culture-enrichment method (ΔC_*T*_=6.69, *p*<0.00001; Figure 2B). Similarly, adjusting for differences in sample storage conditions between the sampling periods, we still observed significantly lower bacterial quantities (as inferred by higher qPCR C_*T*_ values) when using the extraction-free protocol as compared to the culture-enrichment method (ΔC_*T*_=6.68, *p*<0.00001; Supplementary Table 3). This difference in C_*T*_ values between the two methods did not vary by sampling period (ΔC_*T*_=1.26, *p*=0.09; Supplementary Table 4).

**Figure 3.**
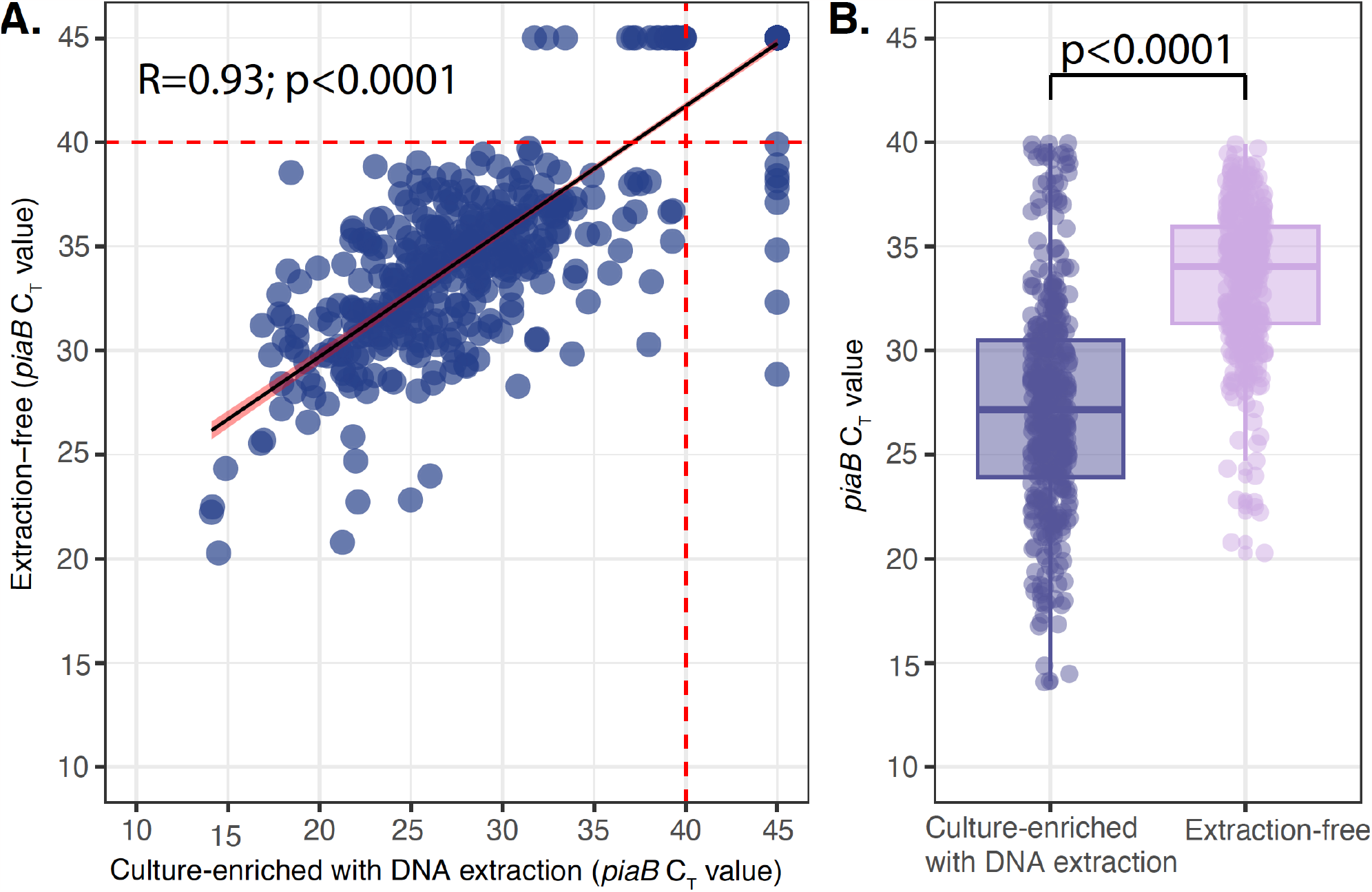
Comparison of piaB qPCR C_T_-values obtained when either lysates from raw saliva or DNA extracted from culture-enriched saliva samples were tested in qPCR for presence of pneumococcus. A) Scatter plot depicting correlation of C_T_ values for pneumococcus-specific gene, piaB, obtained following testing of saliva samples processed by the culture-enrichment and DNA extraction and the extraction-free protocols. The red dotted line marks the threshold assigned to discriminate between positive and negative samples (C_T_-value = 40). B) Box plot of the difference in piaB C_T_ values obtained when saliva samples were tested with the extraction-free method as compared to testing following sample culture-enrichment and DNA extraction. The difference in C_T_-values is depicted on the y-axis, and the average C_T_ value from the samples is on the x-axis.

### Reagent cost for pneumococcal carriage detection per sample

Finally, we conducted a comparison of the expenses associated with laboratory reagents, aiming to assess the cost-effectiveness of the simplified extraction-free method. Overall, the reagent expenses incurred by using saliva lysates (US$2.53/sample) were at least four-fold lower when compared to costs incurred when extracted DNA from culture-enriched samples was used, which ranged from $13.60 to $19.46 depending on whether commercial or in-house made Gent plates were used (Table 2, Supplementary Table 6). The culture-enrichment method took ∼24 hours from sample processing to obtaining a result as it required overnight incubation during the culture-enrichment step (Figure 1), while the extraction-free method only required ∼2 hours to obtain a result. These estimated costs do not account for general laboratory consumables e.g., pipette tips, or required equipment and instruments such as qPCR machines, nor personnel time.

**Table 2.** Costs of pneumococcal carriage detection in saliva samples when tested using qPCR following an extraction-free protocol or culture-enrichment and DNA extraction.

| Items<sup>#</sup> | Price per sample<sup>*</sup> |  |
| --- | --- | --- |
|  | Culture-enrichment with DNA extraction protocol | Extraction-free protocol |
| Saliva collection | \$1.14 | \$1.14 |
| Sample culture-enrichment | \$2.04-\$7.90<sup>1</sup> | - |
| MagMAX™ Viral/Pathogen Nucleic Acid Isolation reagents | \$3.81 | \$0.16<sup>2</sup> |
| Additional consumables for automated DNA extraction on Kingfisher Apex | \$5.38 | - |
| piaB qPCR | \$1.23 | \$1.23 |
| Total cost per sample | \$13.60-\$19.46 | \$2.53 |

<sup>#</sup>*Specific details of reagents, suppliers/brands and their respective catalog numbers are included in Supplementary Table 6.*
<sup>\*</sup>*Price per sample is calculated based on prices listed on the vendor websites and does not include additional costs for general laboratory consumables e.g., pipette tips, or required equipment and instruments such as qPCR machines, nor personnel time.*
<sup>1</sup>*Price range of each sample processed by culture-enrichment depends on whether a commercial or in-house made Gent plate was used.*
<sup>2</sup>*Includes only cost for proteinase K*

## DISCUSSION

Oral samples have emerged as a promising sample type for the surveillance of respiratory pathogens.^30^ For the detection of pneumococcus, the recommended approach for polymicrobial samples such as saliva, is to culture-enrich samples, preferably on blood agar plates supplemented with gentamicin and perform a qPCR on DNA extracted from the culture-enriched samples.^14^ Deviating from this recommended approach could impact the sensitivity of detection, as culture-enrichment increases the pneumococcal load in the sample and the DNA extraction processes concentrates the DNA through its elution into a smaller volume compared to the input volume of saliva. Nonetheless, efforts to utilize minimally processed saliva samples for pneumococcal detection to reduce sample processing time and required resources have been made.^17^ Having developed a saliva-based, extraction-free PCR test for SARS-CoV-2 to simplify testing during the COVID-19 pandemic, we expanded upon this protocol, and evaluated its potential use for the detection of pneumococcus as a more cost-effective and sustainable option for pneumococcal carriage surveillance. Prior to this, limited evidence existed for whether complete removal of the DNA extraction and purification step would provide quality and sufficient DNA template for qPCR amplification, particularly for polymicrobial samples. Yet, remarkably, when we applied a culture- and extraction-free protocol to raw unsupplemented saliva, we obtained comparable rates of pneumococcal detection, to when DNA extracted from culture-enriched saliva was tested. Moreover, the estimated cost of pneumococcal detection using the extraction-free method was three-to-four-fold lower compared to DNA extracted from culture-enriched saliva. This finding suggests that an extraction-free method could provide a less resource-intensive option for carriage detection as compared to the widely utilized culture-enrichment approach.

Despite the overall higher concordance in pneumococcal detection and a strong C_*T*_ values correlation observed between the two methods, C_*T*_ values obtained from the saliva lysates were weaker compared to those obtained from culture-enriched saliva. The observed differences in the resulting C_*T*_ values between the two methods (?C_*T*_ value=6.69) were not unexpected. This observed reduction in sensitivity is in line with previous findings where a difference in C_*T*_ values was observed when comparing DNA extracted from saliva, with and without culture-enrichment.^3^ Collectively, these findings indicate that the observed impact on C_*T*_ values in the current study primarily result from the absence of culture enrichment rather than total omission of DNA extraction using commercial kits, which additionally offers purification and concentration of the target. Given that DNA extraction did not have a significant impact on sensitivity of detection of pneumococcus, our findings support the possibility of omitting DNA extraction using commercial kits when testing raw saliva samples without the culture-enrichment step. With weaker C_*T*_ values obtained when using the extraction-free method suggesting that the use of saliva lysates may have limitations when applied to samples with low pneumococcal bacterial load, the extraction-free method can also be used to identify samples that can be prioritized for culture and DNA extraction, thereby drastically reducing testing costs and time required for detection.

In several instances, we detected discordant results between the extraction-free protocol and culture-enrichment method. The discrepancies were observed mostly in samples with C_*T*_ values >35 from either method. In cases where a positive result was yielded using culture-enriched saliva only, it is likely due to the higher limit of detection of the extraction-free assay, particularly in samples with lower pneumococcal load. The cause for these inconsistent results in cases where the sample tested positive by the extraction-free method only is not entirely clear, but could be driven by possible detection of residual DNA in the absence of live cells or possible growth inhibition of pneumococcus by commensal flora in saliva during the culture-enrichment step, hence yielding a positive result on the extraction-free method but being missed when saliva is culture-enriched.^27^ Although this result was unexpected, it highlights situations where culture-enrichment may underperform in polymicrobial samples and therefore, further studies are required to investigate these situations further to confirm these observations.

A major limitation of our study is that we exclusively used samples collected from young children, a population known for high pneumococcal densities.^28,29^ Therefore, our results cannot be generalized to other age groups particularly the adult population, in which both overall carriage as well as carriage density are typically lower, while non-pneumococcal *Streptococcus* spp. may also be more prevalent.^30^ In this study, we classified samples as positive for pneumococcus using only the *piaB* qPCR assay. While *lytA*^*31*^ and *sp2020*^*32*^ assays are also commonly used, they lack specificity for pneumococcus as compared to *piaB* as closely related non-pneumococcal *Streptococcus* spp. carry homologues of these genes.^30,32^ Therefore, to evaluate our sample processing methods for their impact on pneumococcal carriage detection and to exclude confounding from other streptococci, we limited our analyses to *piaB*.^19,30^ Nonetheless, results obtained here, when testing the culture-enriched saliva samples using *piaB*, highly correlated with results obtained when targeting the *lytA* gene in the same culture-enriched saliva samples (see Supplementary Figure 2). However, a multiplex PCR assay of the key targets used for pneumococcal carriage detection could be further evaluated. Similarly, we did not evaluate the impact of the extraction-free method on pneumococcal serotype detection; confirming its performance when combined with serotype-specific qPCR assays will be important should this method be applied in studies estimating the distribution of vaccine-type pneumococcal prevalence rates to evaluate the impact of novel vaccination strategies for pneumococcus.

In conclusion, our results support the potential use of a culture- and extraction-free protocol as a cost-effective alternative to the more resource-intensive culture-enrichment-based method for pneumococcal carriage detection in children. With broad application, this extraction-free approach could significantly reduce the costs of surveillance efforts. This cost-saving benefit could support either a greater number of studies to be conducted in more regions, or large sample size collection to generate comprehensive data on pneumococcal carriage in these communities. Data generated from these surveillance efforts will provide insight into the effectiveness of the existing pneumococcal vaccines and help to inform future pneumococcal vaccine strategies. Moreover, when combined with already existing saliva-based, extraction-free assays for the detection of respiratory tract viruses, these assays can be used concurrently for the detection of viral-pneumococcal co-infection, allowing us to better understand the relationship between these pathogens and facilitate the implementation of effective clinical and public health responses.

## Data Availability

Data will be available upon reasonable request to the authors

## Acknowledgements

We thank the study participants and their families for their time and dedication to our study. We thank all members of the Trackcare study team, Yale Pathology Labs, and the Wyllie Lab at the Yale School of Public Health for their critical contributions to the initial research study from which these samples were obtained.

## AUTHOR CONTRIBUTIONS

ALW, OMA and CP conceptualized the study. AKB and ALW managed the collection of samples. TL, DYC, KT and OMA developed and validated the laboratory methods. CP, TL and AY conducted laboratory experiments. CP and TL analyzed the data. DMW and VG provided statistical advice. CP, TL and ALW drafted the manuscript. All authors amended, critically reviewed, and commented on the final manuscript.

