## Supplementary Information for "A low-cost culture- and DNA extraction-free method for the molecular detection of pneumococcal carriage in saliva"

*These authors contributed equally

**Supplementary Table 1. qPCR $C_T$ values for samples yielding non-concordant results for pneumococcal detection following testing using the extraction-free method and the culture-enrichment method.**

| Sample ID | Extraction-free method ($C_T$ values) | Culture-enrichment method ($C_T$ values) |
|---|---|---|
| Sample 1 | 45 | 33.42 |
| Sample 2 | 45 | 31.73 |
| Sample 3 | 45 | 32.41 |
| Sample 4 | 45 | 36.87 |
| Sample 5 | 45 | 37.12 |
| Sample 6 | 45 | 37.25 |
| Sample 7 | 45 | 38.04 |
| Sample 8 | 45 | 38.43 |
| Sample 9 | 45 | 38.53 |
| Sample 10 | 45 | 38.93 |
| Sample 11 | 45 | 38.97 |
| Sample 12 | 45 | 39.28 |
| Sample 13 | 45 | 39.4 |
| Sample 14 | 45 | 39.49 |
| Sample 15 | 45 | 39.49 |
| Sample 16 | 45 | 39.49 |
| Sample 17 | 45 | 39.59 |
| Sample 18 | 45 | 39.89 |
| Sample 19 | 45 | 39.92 |
| Sample 20 | 45 | 39.94 |
| Sample 21 | 38.85 | 45 |
| Sample 22 | 32.31 | 45 |
| Sample 23 | 34.83 | 45 |
| Sample 24 | 37.1 | 45 |
| Sample 25 | 37.82 | 45 |
| Sample 26 | 38.15 | 45 |
| Sample 27 | 38.39 | 45 |
| Sample 28 | 38.9 | 45 |
| Sample 29 | 39.89 | 45 |

**Supplementary Table 3. $C_T$ value differences obtained using culture-enrichment and extraction-free methods.**

| | **Δ $C_T$ value** | **Standard Error** | ***p*-value*** |
|---|---|---|---|
| Extraction free method (vs CE method) | +6.69 | 0.311 | <0.00001 |
| School year 2021/2022 | -1.24 | 0.378 | 0.00107 |

**p*-values are derived from a linear regression model

**Supplementary Table 4. $C_T$ value relationship between detection method and sampling season.**

| | **Δ $C_T$ value** | **Standard Error** | ***p*-value*** |
|---|---|---|---|
| Extraction free method (vs CE method) | +5.70 | 0.669 | <0.00001 |
| School year 2021/2022 (vs 2020/2021) | -1.87 | 0.534 | 0.000490 |
| Method: School year interaction | +1.26 | 0.755 | 0.096455 |

**p*-values are derived from a linear regression model

**Supplementary Table 5. Limit of detection for pneumococcus when testing saliva in qPCR following the culture-enrichment and extraction-free sample processing methods.**

| | | DNA extraction-free | | | |
|---|---|---|---|---|---|
| Set number | CFU/ml* | Replicate 1 ($C_T$) | Replicate 2 ($C_T$) | Replicate 3 ($C_T$) | Number of positive samples |
| Rep1 | 5X10^7 | 22.187 | 22.798 | 22.881 | 3/3 |
| Rep1 | 5X10^6 | 25.532 | 26.302 | 26.535 | 3/3 |
| Rep1 | 5X10^5 | 29.202 | 29.947 | 30.312 | 3/3 |
| Rep1 | 5X10^4 | 32.621 | 33.379 | 33.515 | 3/3 |
| Rep1 | 5X10^3 | 36.132 | 37.321 | 35.625 | 3/3 |
| Rep1 | 5X10^2 | 38.800 | ND | 38.816 | 2/3 |
| Rep1 | 5X10^1 | ND[#] | ND | ND | 0/3 |
| Rep2 | 5X10^7 | 23.52 | 23.58 | 23.38 | 3/3 |
| Rep2 | 5X10^6 | 26.66 | 26.88 | 26.8 | 3/3 |
| Rep2 | 5X10^5 | 30.46 | 30.71 | 30.77 | 3/3 |
| Rep2 | 5X10^4 | 33.63 | 33.9 | 33.8 | 3/3 |
| Rep2 | 5X10^3 | 37.31 | 37.86 | 36.96 | 3/3 |
| Rep2 | 5X10^2 | 40.31 | ND | ND | 0/3 |
| Rep2 | 5X10^1 | 40.6 | ND | ND | 0/3 |
| 0 (NEG)[$] | 0 | ND | ND | ND | |
| | | **Culture-enrichment + DNA extraction** | | | |
| Set number | CFU/ml | Replicate 1 | Replicate 2 | Replicate 3 | Number of positive samples |
| Rep1 | 5X10^7 | 16.06 | 15.33 | 15.32 | 3/3 |
| Rep1 | 5X10^6 | 17.93 | 17.36 | 17.75 | 3/3 |
| Rep1 | 5X10^5 | 19.19 | 20.02 | 19.55 | 3/3 |
| Rep1 | 5X10^4 | 22.56 | 24.31 | 22.35 | 3/3 |
| Rep1 | 5X10^3 | 26.99 | 23.92 | 26.44 | 3/3 |
| Rep1 | 5X10^2 | 31.54 | 29.20 | 30.43 | 3/3 |
| Rep1 | 5X10^1 | 36.24 | 34.73 | 31.57 | 3/3 |
| Rep2 | 5X10^7 | 16.20 | 15.92 | 16.02 | 3/3 |
| Rep2 | 5X10^6 | 18.43 | 17.79 | 17.81 | 3/3 |
| Rep2 | 5X10^5 | 20.61 | 21.05 | 21.01 | 3/3 |
| Rep2 | 5X10^4 | 23.24 | 24.56 | 24.96 | 3/3 |
| Rep2 | 5X10^3 | 28.60 | 26.97 | 28.21 | 3/3 |
| Rep2 | 5X10^2 | 31.79 | 31.22 | 33.19 | 3/3 |
| Rep2 | 5X10^1 | ND | 37.61 | 38.50 | 2/3 |
| 0 (NEG) | 0 | ND | ND | ND | |

*CFU = colony forming units; [#]ND = not detectable; [$]NEG = negative control

**Supplementary Table 6. Cost for pneumococcal carriage detection in saliva samples using extraction-free protocol or culture-enrichment and DNA extraction.**

| Items | Supplier/Brand | Catalog Number | Price per sample | | |
|---|---|---|---|---|---|
| | | | Culture-enrichment and DNA extraction | | Extraction-free |
| ***Saliva collection*** | | | | | |
| Falcon tubes (50mL) | Cell Treat | 229435 | $0.37 | | $0.37 |
| Bulb pipette (5 ml) | Fisher Scientific | 13-711-5AM | $0.15 | | $0.15 |
| Cryogenic label | Dymo | | $0.62 | | $0.62 |
| ***Total cost for saliva collection*** | | | ***$1.14*** | | ***$1.14*** |
| ***Saliva culture enrichment*** | | | **Commercial gent plate** | **Inhouse gent plate** | |
| Gent plate | Remel | R01227 | $6.90 | | |
| Spreader | Celltreat | 22961 | $0.54 | $0.54 | |
| Cryovial | Heathrow scientific | HS10060 | $0.28 | $0.28 | |
| Serological pipette | Fisher Sci | 170355 | $0.08 | $0.08 | |
| Brain heart Infusion | BD Diagnostics | 237500 | $0.06 | $0.06 | |
| Glycerol | SigMA-Aldrich | G5516 | $0.04 | $0.04 | |
| Petri dishes | Research Products International | 160268 | | $0.25 | |
| Defibrinated Sheep’s Blood | Colorado Serum Company | 31125 | | $0.62 | |
| TSA II | BD Diagnostics | 212305 | | $0.14 | |
| Gentamicin (10mg/mL) | Life technologies | 15710-064 | | $0.03 | |
| **Total cost for culture-enrichment** | | | **$7.90** | **$2.04** | |
| ***DNA extraction (MagMAX™ Viral/Pathogen Nucleic Acid Isolation Kit)*** | | | | | |
| Magnetic Beads | ThermoFisher | A42362 | $1.25 | | |
| Proteinase K | ThermoFisher | A42363 | $0.16 | | $0.16 |
| Binding Solution | ThermoFisher | A42359 | $0.67 | | |
| Wash Buffer | ThermoFisher | A42360 | $0.73 | | |
| Elution Solution | ThermoFisher | A42364 | $1.00 | | |
| **Total cost for DNA extraction** | | | **$3.81** | | **$0.16** |
| ***Extra items needed when using automated DNA extraction on Apex kingfisher machine*** | | | | | |
| Cryovial tube | Heathrow Scientific | HS10060 | $0.64 | | |
| Sample/wash plates | ThermoFisher | | $2.02 | | |
| Elution plates | ThermoFisher | | $0.38 | | |
| Handystep tips (average) | ThermoFisher | | $2.05 | | |
| 96 well tip comb for Kingfisher Apex | ThermoFisher | | $0.29 | | |
| ***Total for DNA extraction*** | | | ***$5.38*** | | |
| ***S. pneumoniae piaB qPCR*** | | | | | |
| MMX [SSO Advanced] | BioRad | | $0.72 | | $0.72 |
| Molecular grade Water | Research Products International | | $0.01 | | $0.01 |
| Primers | Eurofins | | $0.01 | | $0.01 |
| Probes | Eurofins | | $0.07 | | $0.07 |
| PCR plate | BioRad | | $0.42 | | $0.42 |
| **Total for qPCR detection** | | | **$1.23** | | **$1.23** |
| **Total cost per sample** | | | **$13.60-$19.45** | | **$2.53** |

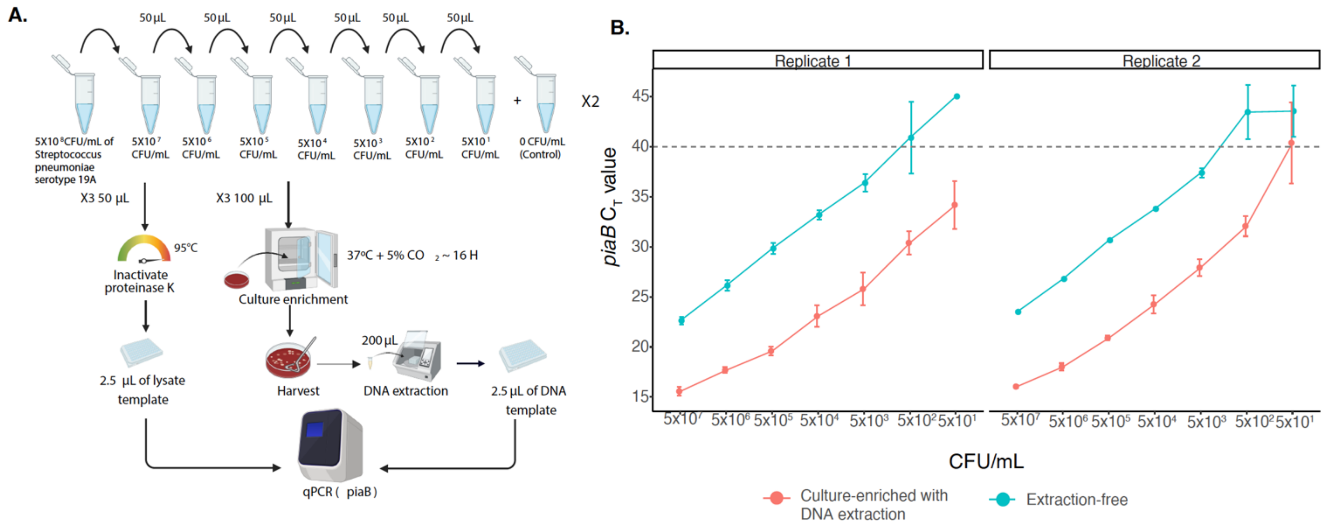


***Supplementary Figure 1. Limit of detection of pneumococcus using extraction-free and culture-enriched methods.*** *A). Workflow used to evaluate the limit of detection for the extraction-free method. B). Limit of assay detection of the extraction-free saliva compared to culture-enriched saliva. Data shown as mean and standard deviation of biological triplicate. CFU = colony forming units, CE = culture-enriched.*

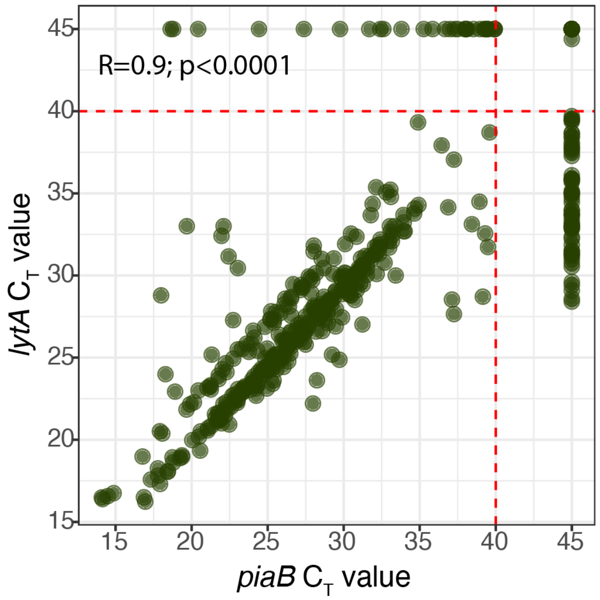


***Supplementary Figure 2. Correlation between qPCR Ct-values obtained when targeting pneumococcal genes, piaB and lytA, when testing DNA templates extracted following culture-enrichment of the saliva samples collected in the study.*** *Scatterplot depicting relationship of $C_T$-values for pneumococcus-specific genes, piaB and lytA, obtained following testing of saliva samples processed by the culture-enrichment. The red dotted line marks the threshold assigned to discriminate between positive and negative samples ($C_T$-value = 40). Correlation coefficient (R) was obtained using the Pearson correlation test.*
